# Education-related inequalities in cause-specific mortality: First estimates for Australia using individual-level linked census and mortality data

**DOI:** 10.1101/2020.09.21.20193516

**Authors:** J Welsh, G Joshy, L Moran, K Soga, HD Law, D Butler, K Bishop, M Gourley, J Eynstone-Hinkins, H Booth, L Moon, N Biddle, A Blakely, E Banks, RJ Korda, for the Whole-of Population Linked Data Project Team

## Abstract

**Background:** Socioeconomic inequalities in mortality are evident in all high-income countries and ongoing monitoring is recommended using linked census-mortality data. Using such data, we provide first estimates of education-related inequalities in cause-specific mortality in Australia, suitable for international comparisons.

**Methods:** Using Australian Census (2016) linked to 13-months of Death Registrations data (2016-17), we estimated relative rates (RR) and rate differences (RD, per100 000 person-years), comparing rates in low (no qualifications) and intermediate (secondary school) with high education (tertiary), for individual causes of death (among those 25-84y) and grouped according to preventability (25-74y), separately by sex and age group, adjusting for age, using negative binomial regression.

**Results:** Among 13.9M people contributing 14 452 732 person-years, 84 743 deaths occurred. We observed inequalities in most causes of death for each age-sex group. Among men aged 25-44y, absolute and relative inequalities (low versus high education) were largest for injuries, e.g. transport accidents (RR=10.1 [95%CI: 5.4-18.7], RD=21.1 [15.9-26.3]). Among those aged 45-64y, inequalities were greatest for chronic diseases, e.g. lung cancer (men RR=6.6 [4.9-8.9], RD=55.6 [51.1-60.1]) and ischaemic heart disease (women RR=5.8 [3.7-9.1], RD=19.2 [17.0-21.5]), with similar patterns for people aged 65-84y. When grouped according to preventability, inequalities were large for causes amenable to behaviour change and medical intervention for all ages and causes amenable to injury prevention among young men.

**Conclusions:** Australian education-related inequalities in mortality are substantial, generally higher than international estimates, and related to preventability. Findings highlight opportunities to reduce them and the potential to improve the health of the population.

Key messages
- Using linked Australian Census (2016) and Death Registrations data (2016-17), we provide the first estimates of education-related inequalities in cause-specific mortality for Australia, broadly suitable for international comparisons.
- Among men aged 25-44 years, inequalities were largest for injuries, with mortality rates among those with low education six-to-ten times that of those with high education. Among the mid- and older-age groups, inequalities were largest for chronic diseases, where mortality rates among those with the lowest education were between two- and seven-times those with the highest education.
- In 2016-17, around half of all deaths for men and one-third of deaths for women aged 25-84 were associated with less than tertiary education. The majority of these excess deaths were attributable to leading causes.
- The substantial inequalities seen in preventable deaths highlight ongoing opportunities to reduce inequalities in mortality and to improve the overall health of the Australian population.
- Australian estimates are generally consistent with, but higher than, those for comparable countries and earlier time periods, but further standardisation of methods and reporting would enhance the validity of such comparisons

## Background

Death rates in high-income countries, including Australia, have decreased substantially over recent decades (**1**), but clear inverse socioeconomic gradients in mortality persist (**2-6**). Understanding the reasons for these inequalities, including identifying causes of death with the largest contribution to these differences, is crucial for informing strategies to reduce health inequalities and improve the overall health of the population. This requires accurate measurement and ongoing monitoring of inequalities in cause-specific mortality, including the ability to compare inequalities across countries and over time.

The OECD recommends measuring inequalities using longitudinal, census-linked-to-mortality data, with education as the socioeconomic indicator (**7**). Many high-income countries, including most European countries, monitor inequalities using this approach and have shown that inequalities vary substantially by cause of death. Consistent with the notion that inequalities reflect unequal distribution of resources required to protect and promote good health, inequalities are larger for causes of death amenable to prevention, including injury, causes linked to smoking and excessive alcohol consumption, and causes amenable to medical care, compared with other causes of death (**8, 9**).

In Australia, routine estimates of inequalities in cause-specific mortality are based on area-level measures of socioeconomic position (SEP). This approach, most commonly using Socio-Economic Indexes for Areas (SEIFA) Index of Relative Socio-Economic Disadvantage (IRSD) quintiles (**10-13**), makes it difficult to compare inequality estimates with other countries. Further, this method misclassifies people in regard to their individual-level SEP because of the heterogeneity within statistical areas on which these measures are based (which contain an average of 400 people) (**14**). This typically results in lower estimates of inequalities compared to those based on individual-level measures (**15**). While individual-level SEP measures are not collected in mortality data in Australia, recent developments in linkage of national data has led to the availability of these data through linkage with census data. Thus, in line with international standards, education-related inequalities in mortality can be quantified.

The aim of this study was to quantify, for the first time, relative and absolute education-related inequalities in cause-specific mortality, including the leading causes of death and causes categorised according to preventability, for Australia using census-linked-to-death data.

## Method

We used linked 2016 Census of Population and Housing and 2016-17 Death Registrations data to create a cohort study of the resident population of Australia, followed-up for 13-months for cause-specific mortality.

### Data sources and sample

Data came from the Multi-Agency Data Integration Project (MADIP), a partnership among Australian Government agencies to link administrative and survey data, including data relating to demographic characteristics and health. Underpinning MADIP data is a Person Linkage Spine, used to create a person-level identification key by linking data from three administrative databases, together resulting in virtually complete coverage of the resident population (**16**): Medicare Enrolments Database (records of those covered by Medicare, Australia’s universal health insurer); Social Security and Related Information database (records of those receiving government benefits); and, Personal Income Tax database (records of those who lodge a tax return). The Spine is the dataset to which all other data sources are linked and contains basic demographic information only. In this study, the 2016 Census was linked with Death Registrations data via the Spine. Linkage was performed using deterministic and probabilistic linking methods, using name, full date of birth, address and sex, with linkage rates of 92% for the Census and 97% for deaths (**16**).

The scope of the 2016 Census was usual residents of Australia on the night of 9 August 2016 living in private and non-private dwellings (**17**). It had an estimated person response rate of 94.8%, with some variation in response by ethnicity and location (**18**). We included all usual residents aged 25-84 years whose census record was linked to the Spine. Death Registrations data contained information on month and year of death occurrence, and underlying cause of death for all deaths registered in Australia in the 2016 and 2017 calendar years (**19**). Death Registrations data were complete until August 2017, allowing for an almost 13-month follow-up period.

### Variables

#### Education

We derived highest level of education from two census variables: highest year of school completed (from ≤Year 8 to Year 12 or equivalent) and highest non-school qualification (from No non-school qualification, to Postgraduate Degree). We created three education categories, corresponding to International Standard Classification of Education (ISCED) categories (**20**): low education (no secondary school graduation or other qualification, ISCED levels 0-2), intermediate education (secondary graduation with/without other non-tertiary qualifications, ISCED levels 3-5) and high education (tertiary qualification, irrespective of secondary school level, ISCED levels 6-8). Missing data on education (5.3%) were imputed using single imputation with ordered logistic regression (Supplementary 1).

#### Cause of death

Underlying cause of death was coded according to the International Classification of Diseases and Related Health Problems, Tenth Revision (ICD-10) and grouped using the Australian Bureau of Statistics method of identifying leading causes of death (**21**) (Supplementary Table 2.1 contains ICD-10 codes). We obtained leading causes directly from the complete Death Registrations file using deaths occurring in the study time period (i.e. August 2016-August 2017). We further grouped causes by broad cause (circulatory diseases, cancers, external causes, infectious diseases and other causes) and preventability, based on established methods, which included causes considered amenable to behaviour change, medical intervention or injury prevention, and non-preventable causes (Supplementary Table 2.2) (**8**). As only month and year of death were available in the Death Registrations file in MADIP, all deaths were assumed to have occurred on the 15^th^ day of the month.

#### Covariates

Age at census, in years, and sex were obtained from the Census.

### Analysis

Prior to our main analysis, we performed data validation analyses to assess potential selection bias from excluding people without a census record and incomplete linkage (Supplementary 3). First, we compared all-cause and cause-specific mortality rates produced using the analysis file with official national estimates and estimates produced using the complete Death Registrations file. Second, we estimated socioeconomic inequalities using SEIFA IRSD (an area-based measure of SEP), comparing estimates produced using the analysis file with those produced using the complete Death Registrations file.

In our main analysis, we estimated relative and absolute education-related inequalities for the 10 leading causes of death for each sex- and age-group, and for causes grouped according to preventability. All analyses were performed separately for men and women and by broad age group.

To quantify relative inequalities in death rates (deaths/person-years), we used negative binomial regression, due to overdispersion in the data, to estimate relative rates (RR) with 95% confidence intervals (CIs) for low and for intermediate compared with high education, focusing on estimates of low vs high education. For each person, person-years-at-risk was the time from the date of the Census (9 August 2016) to the date of death or end of the study period (31 August 2017), whichever occurred first. Analyses were age-adjusted, using 5-year age groups.

To estimate absolute inequalities in death rates, we estimated rate differences per 100 000 (RD), using high education as the reference group. Given absolute death rates were under estimated in our study (Supplementary Tables 3.2-3.3), we maximised external validity of the RDs by estimating the education-specific mortality rates by applying the relevant RRs (described above) to age-sex specific mortality rates for Australia, calculated using data from the complete 2016 Death Registrations file and the 2016 mid-year estimated resident population (**22**). We also estimated the number of annual excess deaths associated with less than tertiary education by multiplying the RDs by the age-sex-specific usual resident population in 2016 with low and intermediate education and summing them.

We also report the relative index of inequality (RII). The RII converts a categorical measure to a continuous measure based on the proportion of people in each education category, and can be interpreted as the ratio of the mortality rates predicted for those on the hypothetical lowest and highest points on the continuous measure (**23**).

In supplementary analyses, we quantified inequalities in broad causes of death, and ranked individual causes by magnitude of relative inequalities, including all leading causes of death and other causes with at least 50 deaths recorded in the analysis file within the relevant age-and sex-group.

Analyses were conducted through the ABS virtual DataLab using Stata 15 (**24**). Ethics approval for this study was granted by the Australian National University Human Research Ethics Committee (reference 2016/666). We notified ABS of this ethics approval as part of a formal application to access the linked dataset in the ABS Virtual Datalab.

## Results

There were 15 562 042 census records for usual residents of Australia aged 25-84 years (**25**) (Figure 1). After excluding records which did not link to the Person Linkage Spine (n=1 700 777, 11%) and records linked in error (n= 603, <0.01%), our final sample included 13 860 662 residents aged 25-84 years (87% of the in-scope population (**22**)), among whom, there were 84 743 deaths (85% of deaths in this age group; 98% of deaths occurring between ages 25-86 years linked to the Spine, Supplementary Table 3.1). After imputation, 26.8% of the sample had low, 47.9% intermediate and 25.3% had high levels of education (Supplementary, Table 1.1).

**Figure 1.**
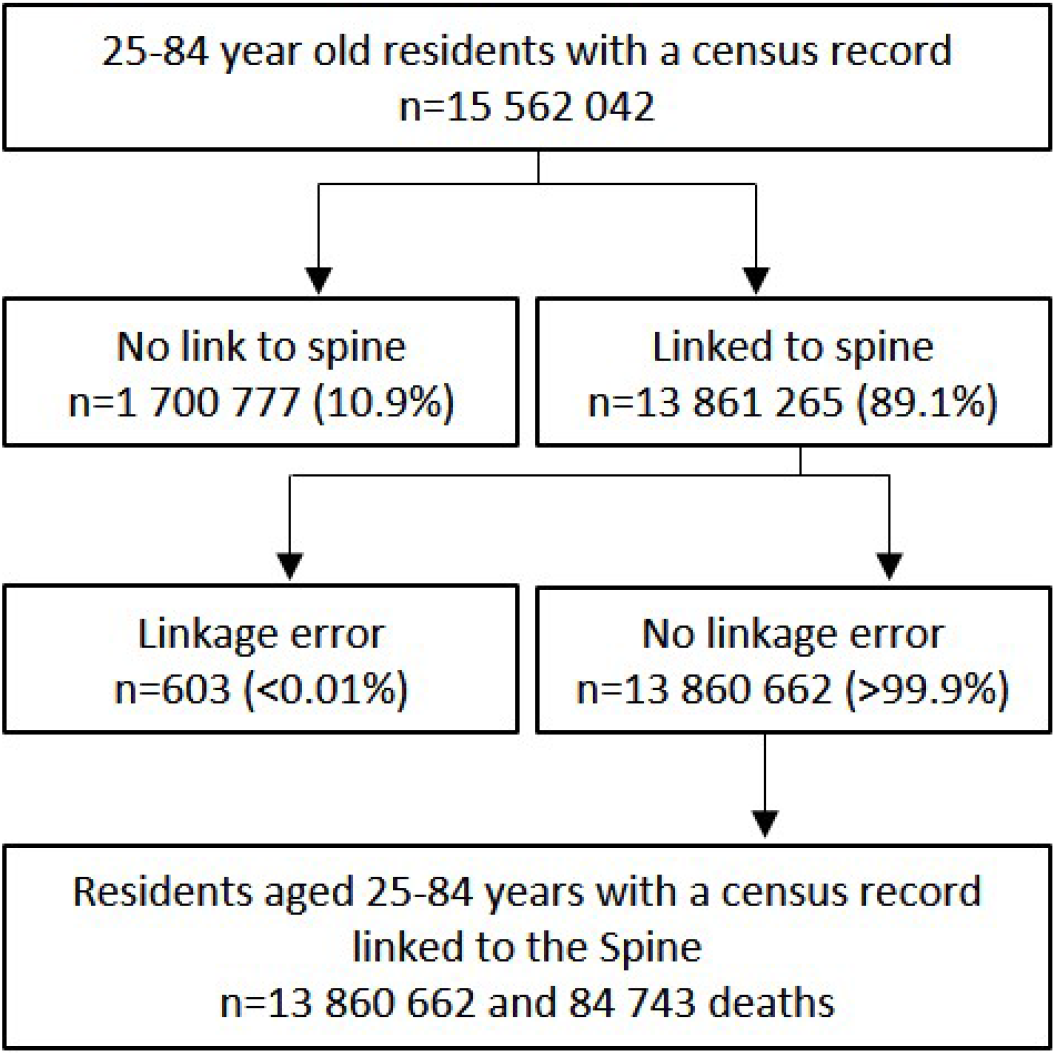
Study flow diagram.

In general, validation analyses provided support for use of the analysis file to quantify education-related inequalities in mortality, as area-level estimates produced using the analysis file were comparable to estimates produced using complete Death Registrations. The exception to this was among younger women where inequality estimates were underestimated the analysis file (Supplementary 3).

### Inequalities in all-cause mortality

All-cause mortality rates were higher in those with lower levels of education: the age-adjusted all-cause mortality RR (low versus high education) was 2.76 (95%CI: 2.61, 2.91) among men aged 25-84 (RD=649 per 100 000 [628, 669]), and 2.13 (2.01, 2.26) among women of the same age (RD=312 per 100 000 [303-320] (Supplementary Tables 4.1-4.2).

### Inequalities in leading causes of death

With few exceptions, those with lower levels of education had higher mortality rates for leading causes of death (Tables 1 and 2). The magnitude of relative and absolute inequalities varied substantially by cause and by age and sex.

**Table 1.**
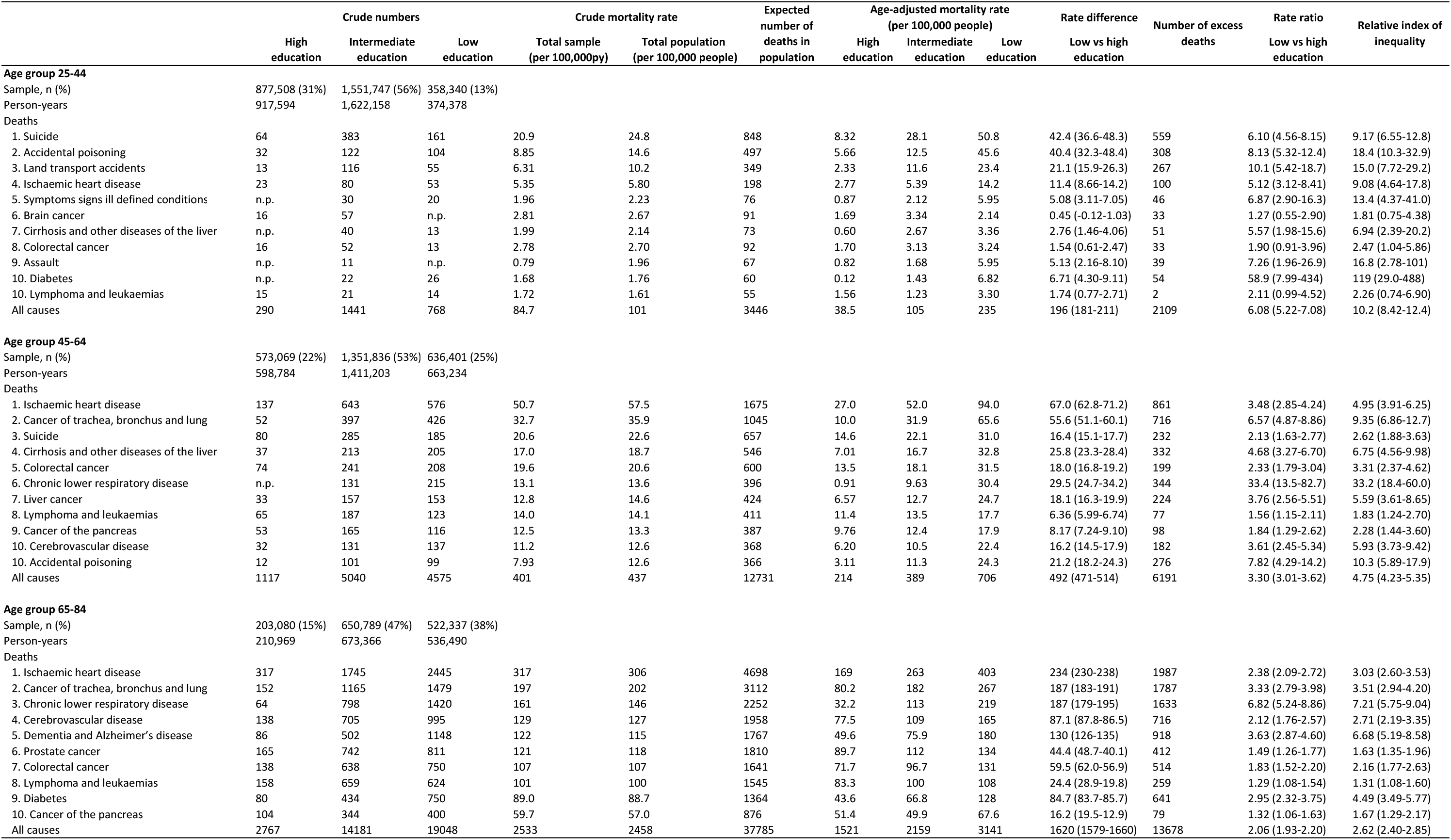
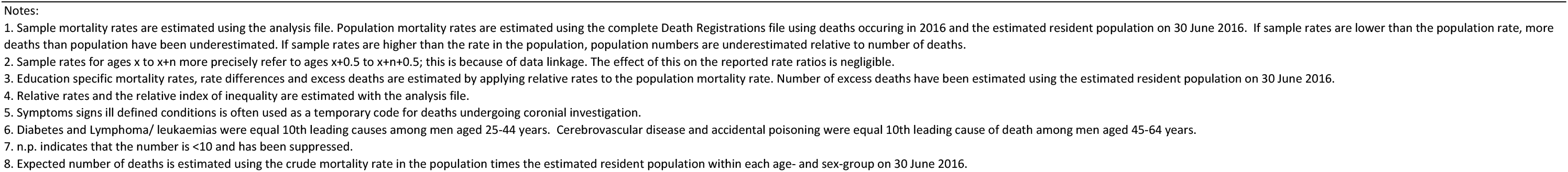
Number of deaths, mortality rates, and absolute and relative inequalities for leading causes of death among Australian men aged 25‐84 years according to education by age group, 2016‐17.

|  | Crude numbers |  |  | Crude mortality rate |  | Expected number of deaths in population | Age-adjusted mortality rate (per 100,000 people) |  |  | Rate difference<br>Low vs high education | Number of excess deaths | Rate ratio<br>Low vs high education | Relative index of inequality |
| --- | --- | --- | --- | --- | --- | --- | --- | --- | --- | --- | --- | --- | --- |
|  | High education | Intermediate education | Low education | Total sample (per 100,000py) | Total population (per 100,000 people) |  | High education | Intermediate education | Low education |  |  |  |  |
| Age group 25-44 |  |  |  |  |  |  |  |  |  |  |  |  |  |
| Sample, n (%) | 877,508 (31%) | 1,551,747 (56%) | 358,340 (13%) |  |  |  |  |  |  |  |  |  |  |
| Person-years | 917,594 | 1,622,158 | 374,378 |  |  |  |  |  |  |  |  |  |  |
| Deaths |  |  |  |  |  |  |  |  |  |  |  |  |  |
| 1. Suicide | 64 | 383 | 161 | 20.9 | 24.8 | 848 | 8.32 | 28.1 | 50.8 | 42.4 (36.6-48.3) | 559 | 6.10 (4.56-8.15) | 9.17 (6.55-12.8) |
| 2. Accidental poisoning | 32 | 122 | 104 | 8.85 | 14.6 | 497 | 5.66 | 12.5 | 45.6 | 40.4 (32.3-48.4) | 308 | 8.13 (5.32-12.4) | 18.4 (10.3-32.9) |
| 3. Land transport accidents | 13 | 116 | 55 | 6.31 | 10.2 | 349 | 2.33 | 11.6 | 23.4 | 21.1 (15.9-26.3) | 267 | 10.1 (5.42-18.7) | 15.0 (7.72-29.2) |
| 4. Ischaemic heart disease | 23 | 80 | 53 | 5.35 | 5.80 | 198 | 2.77 | 5.39 | 14.2 | 11.4 (8.66-14.2) | 100 | 5.12 (3.12-8.41) | 9.08 (4.64-17.8) |
| 5. Symptoms signs ill defined conditions | n.p. | 30 | 20 | 1.96 | 2.23 | 76 | 0.87 | 2.12 | 5.95 | 5.08 (3.11-7.05) | 46 | 6.87 (2.90-16.3) | 13.4 (4.37-41.0) |
| 6. Brain cancer | 16 | 57 | n.p. | 2.81 | 2.67 | 91 | 1.69 | 3.34 | 2.14 | 0.45 (-0.12-1.03) | 33 | 1.27 (0.55-2.90) | 1.81 (0.75-4.38) |
| 7. Cirrhosis and other diseases of the liver | n.p. | 40 | 13 | 1.99 | 2.14 | 73 | 0.60 | 2.67 | 3.36 | 2.76 (1.46-4.06) | 51 | 5.57 (1.98-15.6) | 6.94 (2.39-20.2) |
| 8. Colorectal cancer | 16 | 52 | 13 | 2.78 | 2.70 | 92 | 1.70 | 3.13 | 3.24 | 1.54 (0.61-2.47) | 33 | 1.90 (0.91-3.96) | 2.47 (1.04-5.86) |
| 9. Assault | n.p. | 11 | n.p. | 0.79 | 1.96 | 67 | 0.82 | 1.68 | 5.95 | 5.13 (2.16-8.10) | 39 | 7.26 (1.96-26.9) | 16.8 (2.78-101) |
| 10. Diabetes | n.p. | 22 | 26 | 1.68 | 1.76 | 60 | 0.12 | 1.43 | 6.82 | 6.71 (4.30-9.11) | 54 | 58.9 (7.99-434) | 119 (29.0-488) |
| 10. Lymphoma and leukaemias | 15 | 21 | 14 | 1.72 | 1.61 | 55 | 1.56 | 1.23 | 3.30 | 1.74 (0.77-2.71) | 2 | 2.11 (0.99-4.52) | 2.26 (0.74-6.90) |
| All causes | 290 | 1441 | 768 | 84.7 | 101 | 3446 | 38.5 | 105 | 235 | 196 (181-211) | 2109 | 6.08 (5.22-7.08) | 10.2 (8.42-12.4) |
| Age group 45-64 |  |  |  |  |  |  |  |  |  |  |  |  |  |
| Sample, n (%) | 573,069 (22%) | 1,351,836 (53%) | 636,401 (25%) |  |  |  |  |  |  |  |  |  |  |
| Person-years | 598,784 | 1,411,203 | 663,234 |  |  |  |  |  |  |  |  |  |  |
| Deaths |  |  |  |  |  |  |  |  |  |  |  |  |  |
| 1. Ischaemic heart disease | 137 | 643 | 576 | 50.7 | 57.5 | 1675 | 27.0 | 52.0 | 94.0 | 67.0 (62.8-71.2) | 861 | 3.48 (2.85-4.24) | 4.95 (3.91-6.25) |
| 2. Cancer of trachea, bronchus and lung | 52 | 397 | 426 | 32.7 | 35.9 | 1045 | 10.0 | 31.9 | 65.6 | 55.6 (51.1-60.1) | 716 | 6.57 (4.87-8.86) | 9.35 (6.86-12.7) |
| 3. Suicide | 80 | 285 | 185 | 20.6 | 22.6 | 657 | 14.6 | 22.1 | 31.0 | 16.4 (15.1-17.7) | 232 | 2.13 (1.63-2.77) | 2.62 (1.88-3.63) |
| 4. Cirrhosis and other diseases of the liver | 37 | 213 | 205 | 17.0 | 18.7 | 546 | 7.01 | 16.7 | 32.8 | 25.8 (23.3-28.4) | 332 | 4.68 (3.27-6.70) | 6.75 (4.56-9.98) |
| 5. Colorectal cancer | 74 | 241 | 208 | 19.6 | 20.6 | 600 | 13.5 | 18.1 | 31.5 | 18.0 (16.8-19.2) | 199 | 2.33 (1.79-3.04) | 3.31 (2.37-4.62) |
| 6. Chronic lower respiratory disease | n.p. | 131 | 215 | 13.1 | 13.6 | 396 | 0.91 | 9.63 | 30.4 | 29.5 (24.7-34.2) | 344 | 33.4 (13.5-82.7) | 33.2 (18.4-60.0) |
| 7. Liver cancer | 33 | 157 | 153 | 12.8 | 14.6 | 424 | 6.57 | 12.7 | 24.7 | 18.1 (16.3-19.9) | 224 | 3.76 (2.56-5.51) | 5.59 (3.61-8.65) |
| 8. Lymphoma and leukaemias | 65 | 187 | 123 | 14.0 | 14.1 | 411 | 11.4 | 13.5 | 17.7 | 6.36 (5.99-6.74) | 77 | 1.56 (1.15-2.11) | 1.83 (1.24-2.70) |
| 9. Cancer of the pancreas | 53 | 165 | 116 | 12.5 | 13.3 | 387 | 9.76 | 12.4 | 17.9 | 8.17 (7.24-9.10) | 98 | 1.84 (1.29-2.62) | 2.28 (1.44-3.60) |
| 10. Cerebrovascular disease | 32 | 131 | 137 | 11.2 | 12.6 | 368 | 6.20 | 10.5 | 22.4 | 16.2 (14.5-17.9) | 182 | 3.61 (2.45-5.34) | 5.93 (3.73-9.42) |
| 10. Accidental poisoning | 12 | 101 | 99 | 7.93 | 12.6 | 366 | 3.11 | 11.3 | 24.3 | 21.2 (18.2-24.3) | 276 | 7.82 (4.29-14.2) | 10.3 (5.89-17.9) |
| All causes | 1117 | 5040 | 4575 | 401 | 437 | 12731 | 214 | 389 | 706 | 492 (471-514) | 6191 | 3.30 (3.01-3.62) | 4.75 (4.23-5.35) |
| Age group 65-84 |  |  |  |  |  |  |  |  |  |  |  |  |  |
| Sample, n (%) | 203,080 (15%) | 650,789 (47%) | 522,337 (38%) |  |  |  |  |  |  |  |  |  |  |
| Person-years | 210,969 | 673,366 | 536,490 |  |  |  |  |  |  |  |  |  |  |
| Deaths |  |  |  |  |  |  |  |  |  |  |  |  |  |
| 1. Ischaemic heart disease | 317 | 1745 | 2445 | 317 | 306 | 4698 | 169 | 263 | 403 | 234 (230-238) | 1987 | 2.38 (2.09-2.72) | 3.03 (2.60-3.53) |
| 2. Cancer of trachea, bronchus and lung | 152 | 1165 | 1479 | 197 | 202 | 3112 | 80.2 | 182 | 267 | 187 (183-191) | 1787 | 3.33 (2.79-3.98) | 3.51 (2.94-4.20) |
| 3. Chronic lower respiratory disease | 64 | 798 | 1420 | 161 | 146 | 2252 | 32.2 | 113 | 219 | 187 (179-195) | 1633 | 6.82 (5.24-8.86) | 7.21 (5.75-9.04) |
| 4. Cerebrovascular disease | 138 | 705 | 995 | 129 | 127 | 1958 | 77.5 | 109 | 165 | 87.1 (87.8-86.5) | 716 | 2.12 (1.76-2.57) | 2.71 (2.19-3.35) |
| 5. Dementia and Alzheimer's disease | 86 | 502 | 1148 | 122 | 115 | 1767 | 49.6 | 75.9 | 180 | 130 (126-135) | 918 | 3.63 (2.87-4.60) | 6.68 (5.19-8.58) |
| 6. Prostate cancer | 165 | 742 | 811 | 121 | 118 | 1810 | 89.7 | 112 | 134 | 44.4 (48.7-40.1) | 412 | 1.49 (1.26-1.77) | 1.63 (1.35-1.96) |
| 7. Colorectal cancer | 138 | 638 | 750 | 107 | 107 | 1641 | 71.7 | 96.7 | 131 | 59.5 (62.0-56.9) | 514 | 1.83 (1.52-2.20) | 2.16 (1.77-2.63) |
| 8. Lymphoma and leukaemias | 158 | 659 | 624 | 101 | 100 | 1545 | 83.3 | 100 | 108 | 24.4 (28.9-19.8) | 259 | 1.29 (1.08-1.54) | 1.31 (1.08-1.60) |
| 9. Diabetes | 80 | 434 | 750 | 89.0 | 88.7 | 1364 | 43.6 | 66.8 | 128 | 84.7 (83.7-85.7) | 641 | 2.95 (2.32-3.75) | 4.49 (3.49-5.77) |
| 10. Cancer of the pancreas | 104 | 344 | 400 | 59.7 | 57.0 | 876 | 51.4 | 49.9 | 67.6 | 16.2 (19.5-12.9) | 79 | 1.32 (1.06-1.63) | 1.67 (1.29-2.17) |
| All causes | 2767 | 14181 | 19048 | 2533 | 2458 | 37785 | 1521 | 2159 | 3141 | 1620 (1579-1660) | 13678 | 2.06 (1.93-2.20) | 2.62 (2.40-2.85) |

**Table 2.** Number of deaths, age‐adjusted mortality rates, and absolute and relative inequalities for leading causes of death among Australian women aged 45‐84 years according to education by age group, 2016‐17.

|  | Crude numbers |  |  | Crude mortality rate |  | Expected number of deaths in population | Age-adjusted mortality rate (per 100,000 people) |  |  | Rate difference<br>Low vs high education | Number of excess deaths | Rate ratio<br>Low vs high education | Relative index of inequality |
| --- | --- | --- | --- | --- | --- | --- | --- | --- | --- | --- | --- | --- | --- |
|  | High education | Intermediate education | Low education | Total sample (per 100,000py) | Total population (per 100,000 people) |  | High education | Intermediate education | Low education |  |  |  |  |
| Age group 45-64 |  |  |  |  |  |  |  |  |  |  |  |  |  |
| Sample, n (%) | 671,789 (25%) | 1,211,635 (45%) | 824,955 (30%) |  |  |  |  |  |  |  |  |  |  |
| Person-years | 702,081 | 1,265,756 | 860,976 |  |  |  |  |  |  |  |  |  |  |
| Deaths |  |  |  |  |  |  |  |  |  |  |  |  |  |
| 1. Breast cancer | 168 | 382 | 309 | 30.4 | 30.4 | 921 | 24.9 | 30.9 | 33.9 | 8.94 (8.92-8.97) | 162 | 1.36 (1.12-1.64) | 1.49 (1.16-1.91) |
| 2. Cancer of trachea, bronchus and lung | 72 | 245 | 377 | 24.5 | 25.2 | 762 | 11.4 | 20.8 | 40.0 | 28.6 (26.4-30.8) | 390 | 3.52 (2.68-4.61) | 5.69 (4.05-7.98) |
| 3. Colorectal cancer | 68 | 188 | 160 | 14.7 | 14.3 | 434 | 10.0 | 15.0 | 16.6 | 6.54 (6.34-6.74) | 126 | 1.65 (1.24-2.20) | 1.83 (1.28-2.64) |
| 4. Ischaemic heart disease | 24 | 110 | 204 | 12.0 | 13.0 | 395 | 3.98 | 9.82 | 23.2 | 19.2 (17.0-21.5) | 255 | 5.83 (3.74-9.08) | 10.76 (6.46-17.9) |
| 5. Chronic lower respiratory disease | 11 | 85 | 252 | 12.3 | 10.7 | 323 | 1.48 | 5.98 | 22.2 | 20.7 (18.0-23.4) | 251 | 15.0 (8.06-28.0) | 36.6 (20.1-66.9) |
| 6. Suicide | 32 | 108 | 85 | 7.95 | 7.83 | 237 | 4.43 | 8.26 | 10.1 | 5.66 (4.95-6.37) | 103 | 2.28 (1.49-3.48) | 2.73 (1.60-4.64) |
| 7. Ovarian cancer | 66 | 101 | 97 | 9.33 | 7.76 | 235 | 8.36 | 6.91 | 8.44 | 0.08 (-0.25-0.41) | -19 | 1.01 (0.74-1.38) | 1.07 (0.68-1.67) |
| 8. Cerebrovascular disease | 34 | 77 | 134 | 8.66 | 9.51 | 288 | 5.46 | 6.78 | 16.4 | 10.9 (9.9-11.9) | 119 | 3.00 (2.05-4.38) | 5.82 (3.48-9.75) |
| 9. Cancer of the pancreas | 43 | 99 | 103 | 8.66 | 8.02 | 243 | 6.07 | 7.53 | 9.99 | 3.92 (3.76-4.07) | 56 | 1.64 (1.14-2.36) | 2.01 (1.24-3.26) |
| 10. Cirrhosis and other diseases of the liver | 18 | 70 | 112 | 7.07 | 7.83 | 237 | 2.92 | 6.18 | 13.9 | 11.0 (9.55-12.5) | 145 | 4.78 (2.86-7.99) | 8.70 (4.73-16.0) |
| All causes | 995 | 2804 | 3519 | 259 | 262 | 7927 | 154.9 | 230.3 | 377 | 222 (213-232) | 3058 | 2.44 (2.21-2.68) | 3.46 (3.04-3.94) |
| Age group 65-84 |  |  |  |  |  |  |  |  |  |  |  |  |  |
| Sample, n (%) | 185,930 (12%) | 465,778 (31%) | 856,466 (57%) |  |  |  |  |  |  |  |  |  |  |
| Person-years | 193,558 | 483,988 | 886,110 |  |  |  |  |  |  |  |  |  |  |
| Deaths |  |  |  |  |  |  |  |  |  |  |  |  |  |
| 1. Ischaemic heart disease | 106 | 486 | 1695 | 146 | 140 | 2311 | 68.1 | 109 | 164 | 96.2 (94.2-98.2) | 1092 | 2.41 (1.96-2.97) | 3.07 (2.45-3.85) |
| 2. Dementia and Alzheimer's disease | 96 | 397 | 1538 | 130 | 123 | 2030 | 69.2 | 90.4 | 147 | 78.2 (76.9-79.4) | 829 | 2.13 (1.69-2.68) | 3.11 (2.37-4.08) |
| 3. Chronic lower respiratory disease | 69 | 386 | 1526 | 127 | 118 | 1945 | 40.0 | 80.7 | 146 | 106 (105-108) | 1188 | 3.66 (2.84-4.72) | 5.01 (3.87-6.48) |
| 4. Cancer of trachea, bronchus and lung | 131 | 495 | 1275 | 122 | 124 | 2042 | 75.0 | 109 | 139 | 63.8 (60.3-67.2) | 759 | 1.85 (1.52-2.25) | 2.10 (1.68-2.62) |
| 5. Cerebrovascular disease | 92 | 409 | 1292 | 115 | 111 | 1841 | 62.8 | 94.7 | 127 | 63.9 (61.1-66.8) | 750 | 2.02 (1.60-2.54) | 2.35 (1.82-3.04) |
| 6. Breast cancer | 125 | 342 | 759 | 78.4 | 78.3 | 1294 | 70.6 | 73.6 | 82.4 | 11.7 (5.19-18.30) | 124 | 1.17 (0.96-1.41) | 1.28 (1.02-1.61) |
| 7. Colorectal cancer | 96 | 289 | 733 | 71.5 | 73.8 | 1220 | 61.4 | 67.1 | 79.5 | 18.1 (11.9-24.2) | 195 | 1.29 (1.04-1.61) | 1.48 (1.14-1.92) |
| 8. Lymphoma and leukaemias | 85 | 280 | 596 | 61.5 | 59.3 | 981 | 50.1 | 60.2 | 60.9 | 10.8 (5.36-16.3) | 150 | 1.22 (0.96-1.54) | 1.17 (0.89-1.55) |
| 9. Diabetes | 44 | 144 | 641 | 53.0 | 54.4 | 900 | 29.5 | 33.9 | 66.8 | 37.2 (35.8-38.7) | 367 | 2.26 (1.63-3.14) | 4.08 (2.77-6.01) |
| 10. Cancer of the pancreas | 76 | 203 | 490 | 49.2 | 48.2 | 797 | 43.3 | 43.4 | 51.6 | 8.37 (3.34-13.40) | 78 | 1.19 (0.93-1.53) | 1.39 (1.03-1.88) |
| All causes | 1741 | 6217 | 18780 | 1710 | 1662 | 27474 | 1105 | 1394 | 1866 | 760 (748-774) | 8482 | 1.69 (1.56-1.83) | 2.13 (1.91-2.38) |

**Notes:**

For men aged 25-44 years, relative and absolute inequalities were largest for external causes of death, although there was considerable uncertainty in the estimates due to small numbers of deaths (total deaths=2499, Table 1). This included deaths from land transport accidents, accidental poisoning and suicide (RRs ranged from 6.10 to 10.1, RDs from 21.1 to 42.4 per 100 000, Table 1). Relative inequalities among younger men were also large for ischaemic heart disease and cirrhosis of the liver (RRs were between 5 and 6), although absolute inequalities were small (i.e. <12 per 100 000). There was little evidence of inequalities for brain cancer and colorectal cancer among young men. Due to concerns regarding internal validity, inequality estimates for women aged 25-44 are presented as supplementary material only and should be interpreted with caution (Supplementary Table 3.5).

For men and women aged 45-64 years, relative and absolute and inequalities were largest for cancer of the trachea, bronchus and lung, ischaemic heart disease, cerebrovascular disease, cirrhosis and other liver diseases, and chronic lower respiratory disease (RRs for these causes ranged from 3.00 to 33.4; RDs from 11.0 to 67.0 per 100 000) (Tables 1 and 2). Relative inequalities were also substantial for colorectal cancer, cancer of the pancreas and lymphoma and leukaemias for men and breast cancer for women, although absolute differences were smaller relative to other causes.

In the 65-84 year old age group also, the largest relative and absolute inequalities were observed in chronic diseases (Tables 1 and 2). This included chronic lower respiratory disease, cancer of the trachea, bronchus and lung, ischaemic heart disease, cerebrovascular disease and diabetes (RRs ranged from 1.85 to 6.82, RDs from 37.2 to 234 per 100 000). Among people of this age, absolute and relative inequalities in dementia and Alzheimer’s disease were also considerable.

Among men, the estimated number of excess deaths associated with less than tertiary education from all-causes in the one-year period (2016-17) was 2109 for those aged 25-44, 6191 for those 45-64 and 13 678 among those aged 65-84, equivalent to 61%, 49% and 36% of all deaths in each age group, respectively (Table 1). The estimated number of excess deaths from the 10 leading causes accounted for 71%, 57% and 65% of all excess deaths for those aged 25-44 years, 45-64 years and 65-84 years, respectively. Among women, the number of excess deaths from all causes was 3058 for those aged 45-64 and 8482 for those 65-84, equivalent to 39% and 31% of all deaths among the two age groups, respectively (Table 2). The estimated number of excess deaths from the 10 leading causes was 1589 (52% of the total number of excess deaths from all-causes) for women aged 45-64 and 5533 (65%) among women aged 65-84.

### Inequalities in causes according to preventability

For men and women in each age group, relative inequalities were largest for causes of death amenable to behaviour change (Figure 2, Supplementary Tables 4.3-4.4); absolute inequalities were also generally largest for deaths amenable to behaviour change, with the exception of younger men, where absolute inequalities were largest for causes amenable to injury prevention. That inequalities were generally larger for preventable causes was also evident when all causes of death were ranked by the magnitude of relative inequalities (Supplementary Tables 4.5-4.6). However, small numbers of deaths limited the precision of estimates for some causes.

**Figure 2.**
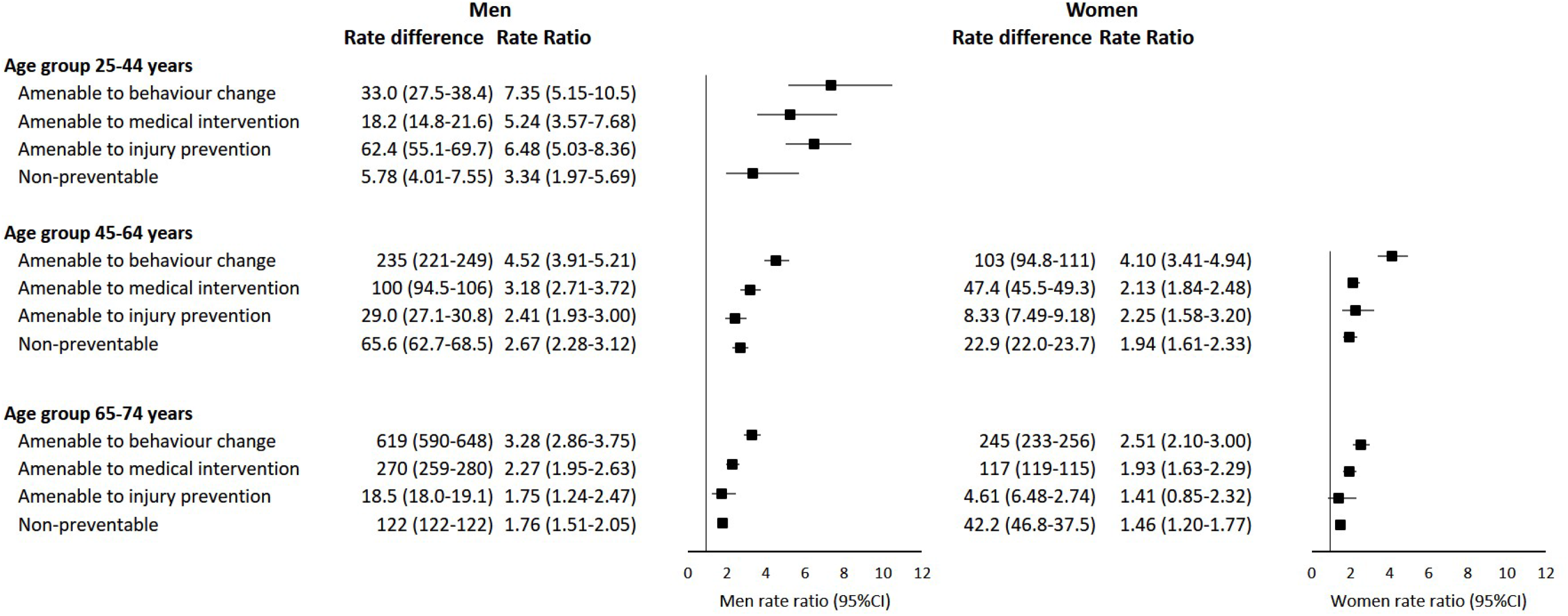
Absolute (per 100 000) and relative inequalities by education (low versus high) for causes of death according to preventability among men aged 25-74 years and women aged 45-74 years by age group, Australia 2016-17. Notes: Rate ratio is plotted. Number of deaths, excess deaths and RIIs for all age-sex groups are available in Supplementary Tables 4.3-4.4. Results are not presented here for women aged 25-44 years due to concerns about the internal validity of the data for this group. They are available in Supplementary Table 3.5 but should be interpreted with caution.

## Discussion

We observed substantial education-related inequalities in virtually all causes of death among the resident population of Australia aged 25-84 years. Among younger men, absolute and relative inequalities were largest for injuries, where mortality rates among those with no educational qualifications were between six (suicide) and 10 (land transport accidents) times that of people with a tertiary education. Among mid-and older-aged men and women, relative and absolute inequalities were largest for chronic diseases, particularly for smoking-related causes, where mortality rates among those with the lowest education were between two- and seven-fold those with the highest education, and two to four-fold for cardiovascular diseases. As expected, relative and absolute inequalities were generally larger for preventable compared to non-preventable causes, and were large for causes amenable to behaviour change and medical intervention across all age groups, and for causes amenable to injury prevention among young men.

This study is the first to comprehensively report on cause-specific education-related inequalities in Australia. Compared with the most recent national estimates of inequalities (for the period 2009-11) using area-level measures of SEP, our education-based inequality estimates are substantially larger for all-cause and cause-specific mortality (**11**). Our estimates are also higher than but consistent with previous estimates of education-related inequalities reported for all-cause (**6**) and selected causes of death (**26**) in Australia for 2011-12. These differences likely reflect, at last in part, methodological differences, as well as changes in the composition of educational groups over time (Supplementary 5).

Our inequality findings for Australia are broadly consistent with those reported for other countries, including the finding that inequalities are larger for preventable compared to non-preventable deaths (**8, 9, 27-29**). However, it is somewhat difficult to directly compare findings across countries. Few cause-specific inequalities studies report RIIs, despite being the recommended method for international comparisons (**7**). Our cause-specific RIIs are consistent with but higher than comparable, earlier, estimates for Italy (2011-12) (**27**), France (1990-1999) (**30**) and Colombia (1998-13 2007) (**29**). The RRs presented in this study are generally larger in magnitude than those reported for other high-income countries (**8, 9, 27-29**). However, similar estimates have been reported in other advanced welfare states (**6, 2, 7, 28**). In Norway, education-related mortality RRs for men in causes amenable to behaviour change were greater than three for the population aged 30-79, and were greater than two for causes amenable to medical intervention (**8**).

The fact that the RRs observed in this study are generally larger than observed in some comparable countries may reflect, at least in part, a greater concentration of disadvantage among those with lower levels of education in Australia (**3, 1, 32**) and/or larger socioeconomic differences in risk factors in Australia compared to other countries. For example, while the proportion of the population who report daily smoking in Australia (12%) is lower compared to many other countries (OECD average, 18%), socioeconomic differences in smoking appear more pronounced (high versus low education: Australia: an absolute difference of 17% (**33**); OECD average: 7% difference (**34**)). Differences in inequality estimates may also reflect methodological and reporting differences rather than true differences in inequalities. In addition to recommendations for standardised methods, recommendations for standardised reporting of inequalities, including sex-age-stratified RIIs, may aid international comparisons and ongoing monitoring of inequalities over time, although differences in linkage methodologies and data quality may continue to limit comparisons. Understanding the mechanisms by which education-related inequalities in mortality occur is critical to ensure that policies are implemented to mitigate them. It was not possible with the data used in this paper to examine specific mechanisms or solutions to reduce inequalities in Australia, but our findings provide insights on areas to target. Among the younger age group, inequalities were largest for external causes of deaths and causes amenable to injury prevention. Although the number of deaths in the younger age group was low, the considerable absolute inequalities in injury-related deaths highlights the potential for further reductions. Among the older age groups, inequalities were greatest for chronic diseases, particularly for causes associated with smoking and alcohol/ substance use. Virtually all behaviour-related risk factors are more prevalent among those of lower compared with higher SEP in Australia (**33**). Our findings further underscore the need for interventions to reduce the disproportionately high prevalence of risk factors among those of lower SEP, including strategies which recognise and address the upstream determinants of these risk factors. We also observed substantial inequalities in cause-specific mortality amenable to medical intervention. This included cancers amenable to screening and diseases amenable to acute medical care, such as cardiovascular diseases. (**35**) While not all deaths in this group of causes could have been avoided with better health care, inequalities in health care are well documented in Australia (e.g. (**36-38**)), and addressing them is likely part of the solution.

Using linked Census and Death Registrations data we had information on 87% of the population of interest, with virtually complete (98%) ascertainment of deaths among those in the sample. Given this, we did not apply a weighting strategy. This is likely to be a valid approach for the middle- and older-age groups, where our mortality rates compared favourably to estimates from the complete population. However, weights may have improved absolute estimates for younger age groups, where rates in the sample were up to 60% lower for some causes relative to the full population. While we addressed this issue by generating absolute inequality estimates by applying RRs to the external population-based mortality rates estimated using the complete Death Registrations file, this method relies on the strong assumption that the RRs are internally valid. This assumption was supported, at least to some degree, by the fact that our study had virtually complete death data among those within the sample and, with the exception of women aged 25-44 years, that there was little evidence that ascertainment of death data differed in relation to area-level SEP. However, given our validation relied on an area-level measure of SEP, we cannot exclude the possibility that linking of death data was differential with respect to education, which would bias our estimates. Furthermore, it was possible that inclusion in to the study was related to both education (exposure) and ill-health (leading to death, the outcome) which would bias our estimates. Given high population coverage, it is likely that the effect of any such selection bias is minimal. We measured mortality occurring over a 13-month follow-up period resulting in small numbers of deaths, particularly for younger age groups and for less common causes of death, limiting the precision of some of our estimates. Longer follow-up periods may be needed for more reliable estimates. Furthermore, delays in death registrations may have contributed to lower mortality rates among younger age groups, which may be improved with updated data. Finally, we did not account for migration or deaths occurring outside of Australia. Given our relatively short follow-up period, it is unlikely that this had a material effect on our estimates.

## Conclusions

Using linked census mortality data enabled valid estimates of education-related inequalities in mortality in Australia, broadly suitable for international comparisons. Standardising the reporting of census-mortality analyses would further enhance the ability to compare estimates across time and countries, although differences in linkage methods and data quality may continue to impede comparisons.

Education-related inequalities are substantial in Australia and evident for most causes of death. The absolute and relative inequalities are largest for preventable deaths, in particular deaths due to injury in younger adults and deaths from preventable cancers and cardiovascular diseases among middle- and older-aged adults. These findings highlight opportunities to reduce health inequalities in Australia and the marked potential to improve the overall health of the population.

## Data Availability

Data from the Multi-Agency Data Integration Project are available for approved projects to approved government and non-government users.

https://www.abs.gov.au/websitedbs/D3310114.nsf/home/Statistical+Data+Integration+-+MADIP

## Data availability statement

Data part of the Multi-Agency Data Integration Project are available for approved projects to approved government and non-government users. https://www.abs.gov.au/websitedbs/D3310114.nsf/home/Statistical+Data+Integration+-+MADIP

## Acknowledgements

We acknowledge the contributions of members of the Whole-of Population Linked Data Project team, including Chief Investigators: Walter Abhayaratna, Bianca Calabria, Louisa Jorm, Raymond Lovett, John Lynch, Naomi Priest; Associate Investigator Rosemary Knight; Partner Investigators: Louise Gates, Gary Jennings, Talei Parker, Clare Saunders, Bill Stavreski; and Project Manager: Katie Beckwith. We also thank Gwenda Thompson and James Chipperfield for their assistance.

---

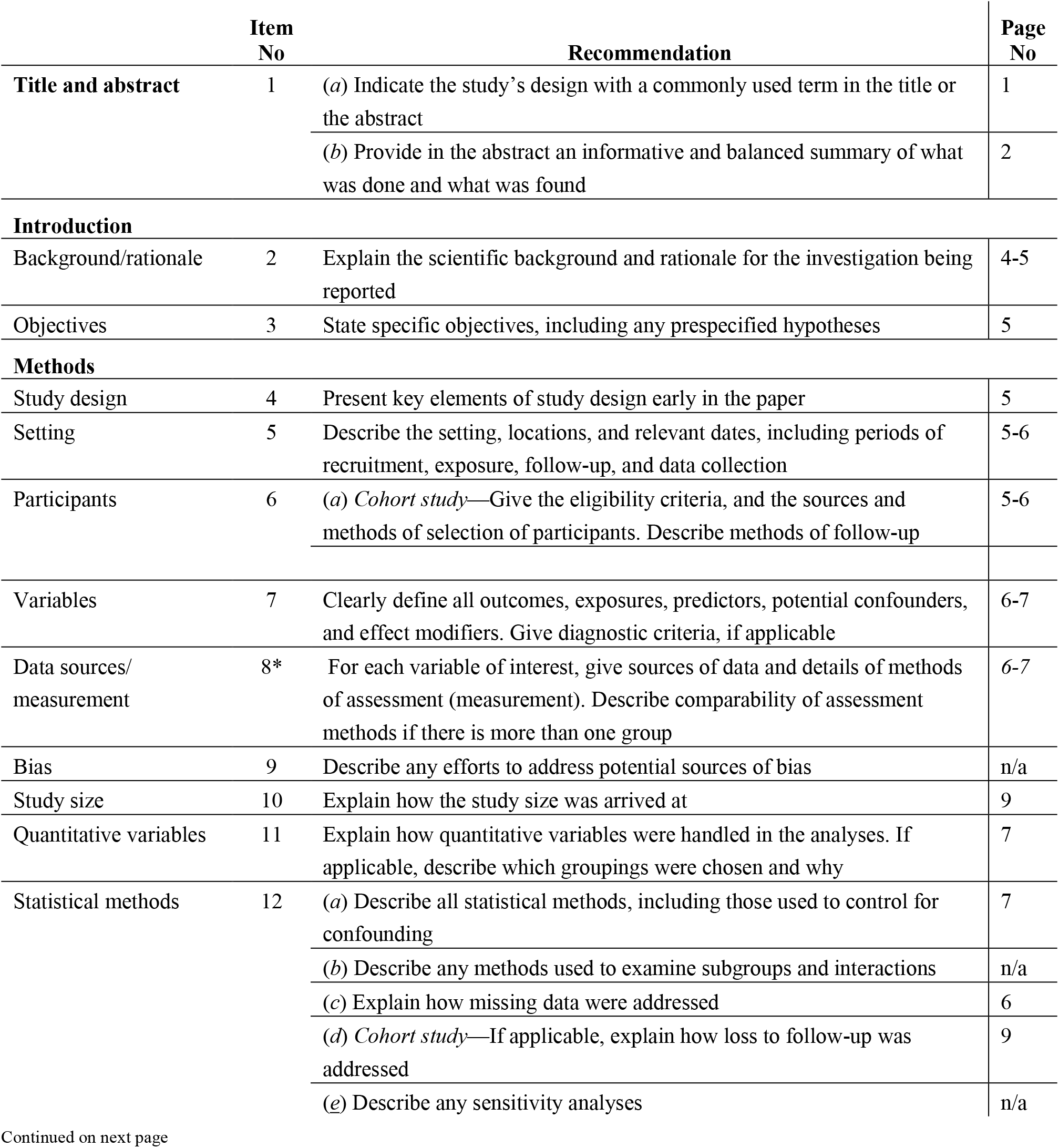
STROBE Statement—checklist of items that should be included in reports of observational studies

|  | Item No | Recommendation | Page No |
| --- | --- | --- | --- |
| **Title and abstract** | 1 | (a) Indicate the study's design with a commonly used term in the title or the abstract | 1 |
|  |  | (b) Provide in the abstract an informative and balanced summary of what was done and what was found | 2 |
| **Introduction** |  |  |  |
| Background/rationale | 2 | Explain the scientific background and rationale for the investigation being reported | 4-5 |
| Objectives | 3 | State specific objectives, including any prespecified hypotheses | 5 |
| **Methods** |  |  |  |
| Study design | 4 | Present key elements of study design early in the paper | 5 |
| Setting | 5 | Describe the setting, locations, and relevant dates, including periods of recruitment, exposure, follow-up, and data collection | 5-6 |
| Participants | 6 | (a) *Cohort study*—Give the eligibility criteria, and the sources and methods of selection of participants. Describe methods of follow-up | 5-6 |
| Variables | 7 | Clearly define all outcomes, exposures, predictors, potential confounders, and effect modifiers. Give diagnostic criteria, if applicable | 6-7 |
| Data sources/<br>measurement | 8* | For each variable of interest, give sources of data and details of methods of assessment (measurement). Describe comparability of assessment methods if there is more than one group | 6-7 |
| Bias | 9 | Describe any efforts to address potential sources of bias | n/a |
| Study size | 10 | Explain how the study size was arrived at | 9 |
| Quantitative variables | 11 | Explain how quantitative variables were handled in the analyses. If applicable, describe which groupings were chosen and why | 7 |
| Statistical methods | 12 | (a) Describe all statistical methods, including those used to control for confounding | 7 |
|  |  | (b) Describe any methods used to examine subgroups and interactions | n/a |
|  |  | (c) Explain how missing data were addressed | 6 |
|  |  | (d) *Cohort study*—If applicable, explain how loss to follow-up was addressed | 9 |
|  |  | (e) Describe any sensitivity analyses | n/a |

Continued on next page

|  |  |  |  |
| --- | --- | --- | --- |
| **Results** |  |  |  |
| Participants | 13* | (a) Report numbers of individuals at each stage of study—eg numbers potentially eligible, examined for eligibility, confirmed eligible, included in the study, completing follow-up, and analysed | 9 |
|  |  | (b) Give reasons for non-participation at each stage | 9 |
|  |  | (c) Consider use of a flow diagram | 9 |
| Descriptive data | 14* | (a) Give characteristics of study participants (eg demographic, clinical, social) and information on exposures and potential confounders | 9 |
|  |  | (b) Indicate number of participants with missing data for each variable of interest | 6 |
|  |  | (c) *Cohort study*—Summarise follow-up time (eg, average and total amount) | 7 |
| Outcome data | 15* | *Cohort study*—Report numbers of outcome events or summary measures over time | 9 |
| Main results | 16 | (a) Give unadjusted estimates and, if applicable, confounder-adjusted estimates and their precision (eg, 95% confidence interval). Make clear which confounders were adjusted for and why they were included | T1, T2 |
|  |  | (b) Report category boundaries when continuous variables were categorized | n/a |
|  |  | (c) If relevant, consider translating estimates of relative risk into absolute risk for a meaningful time period | T1, T2 |
| Other analyses | 17 | Report other analyses done—eg analyses of subgroups and interactions, and sensitivity analyses | 11 |
| **Discussion** |  |  |  |
| Key results | 18 | Summarise key results with reference to study objectives | 11-12 |
| Limitations | 19 | Discuss limitations of the study, taking into account sources of potential bias or imprecision. Discuss both direction and magnitude of any potential bias | 14 |
| Interpretation | 20 | Give a cautious overall interpretation of results considering objectives, limitations, multiplicity of analyses, results from similar studies, and other relevant evidence | 15 |
| Generalisability | 21 | Discuss the generalisability (external validity) of the study results | 14 |
| **Other information** |  |  |  |
| Funding | 22 | Give the source of funding and the role of the funders for the present study and, if applicable, for the original study on which the present article is based | 15 |

\*Give information separately for cases and controls in case-control studies and, if applicable, for exposed and unexposed groups in cohort and cross-sectional studies.
**Note:** An Explanation and Elaboration article discusses each checklist item and gives methodological background and published examples of transparent reporting. The STROBE checklist is best used in conjunction with this article (freely available on the Web sites of PLoS Medicine at http://www.plosmedicine.org/, Annals of Internal Medicine at http://www.annals.org/, and Epidemiology at http://www.epidem.com/). Information on the STROBE Initiative is available at www.strobe-statement.org.

